# A quantitative home-use framework for assessing fertility and identifying novel hormone trends by recording urine hormones

**DOI:** 10.1101/2021.05.11.21257023

**Authors:** Siddharth Pattnaik, Dipankar Das, Varun Akur Venkatesan

## Abstract

Fertility testing using urinary hormones has been used to effectively improve the likelihood of pregnancy. To provide fertility scores, the existing home-use systems measure one or two hormones. However, the hormone profiles vary depending on cycle duration, fertility-related disorders, drugs and other treatments. Here, we introduce Inito, a mobile-phone connected reader that is scalable to multiple hormone tests. In this report, we assess the accuracy of the quantitative home-based fertility monitor, the Inito Fertility Monitor (IFM), and suggest using IFM as a device to monitor and analyse female hormone patterns. There were two aspects of the study: i. evaluation of the efficacy of IFM in quantifying urinary Estrone-3-glucuronide (E3G), pregnanediol glucuronide (PdG) and luteinizing hormone (LH), and ii. A retrospective study of patients’ hormone profiles using IFM. We observed that with all three hormones, IFM had an accurate recovery percentage and had a CV of less than 10 percent. Furthermore, in predicting the concentration of urinary hormones, IFM showed a high correlation with ELISA. Using Inito in clinical trials, we report a novel criterion for earlier confirmation of ovulation compared to existing criteria. We also present a novel hormone pattern consistent across most menstrual cycles included in the study. In conclusion, the Inito Fertility Monitor is an effective tool for calculating the urinary concentrations of E3G, PdG and LH and can also be used to provide accurate fertility scores and confirm ovulation. In addition, the sensitivity of IFM facilitates the monitoring of menstrual cycle-related hormone patterns, therefore also making it a great tool for physicians to track the hormones of their patients.

## Introduction

For couples seeking to conceive, the timing of intercourse is a critical question. Almost 45-50 percent of women do not know their fertile window^1,2^. Calendar-based methods cannot accurately predict the fertile window. During the fertile window, having intercourse increases the likelihood of becoming pregnant^3^. Based on different factors, such as age, diet, genetic predisposition and any infertility disorder, the fertile window may vary.

The pituitary hormones FSH and LH and steroid hormones estrogen and progesterone regulate follicle formation, egg release and uterine preparation for implantation. The predictable behavior of these hormones in a regular menstrual cycle can be exploited to forecast fertile days (Figure 1a). Lateral flow assay-based luteinizing hormone tests at home have widely been used to estimate fertile days. These tests, however, provide a window of 1-2 fertile days(s) near the ovulation day. Previous studies have concluded that the fertile window usually lasts for 6 days and involves the ovulation date^4–6^. Measuring estrone-3-glucuronide (E3G) along with LH can potentially boost the fertility window from 2 days to 6 days. Predicting fertility status by evaluating the levels of LH and oestrogen is also highly correlated with transvaginal ultrasound ovulation prediction^7^. Fertile window estimation, however, is not a guarantee of ovulation. In fact, approximately 26-37% of natural cycles are anovulatory^8^. This may lead to frustration for couples trying to conceive. Therefore, the confirmation of ovulation is sought after by both clinicians and patients, and an additional measurement of pregnanediol-3-glucuronide (PdG) can be used to confirm ovulation^9,10^.

**Figure 1:**
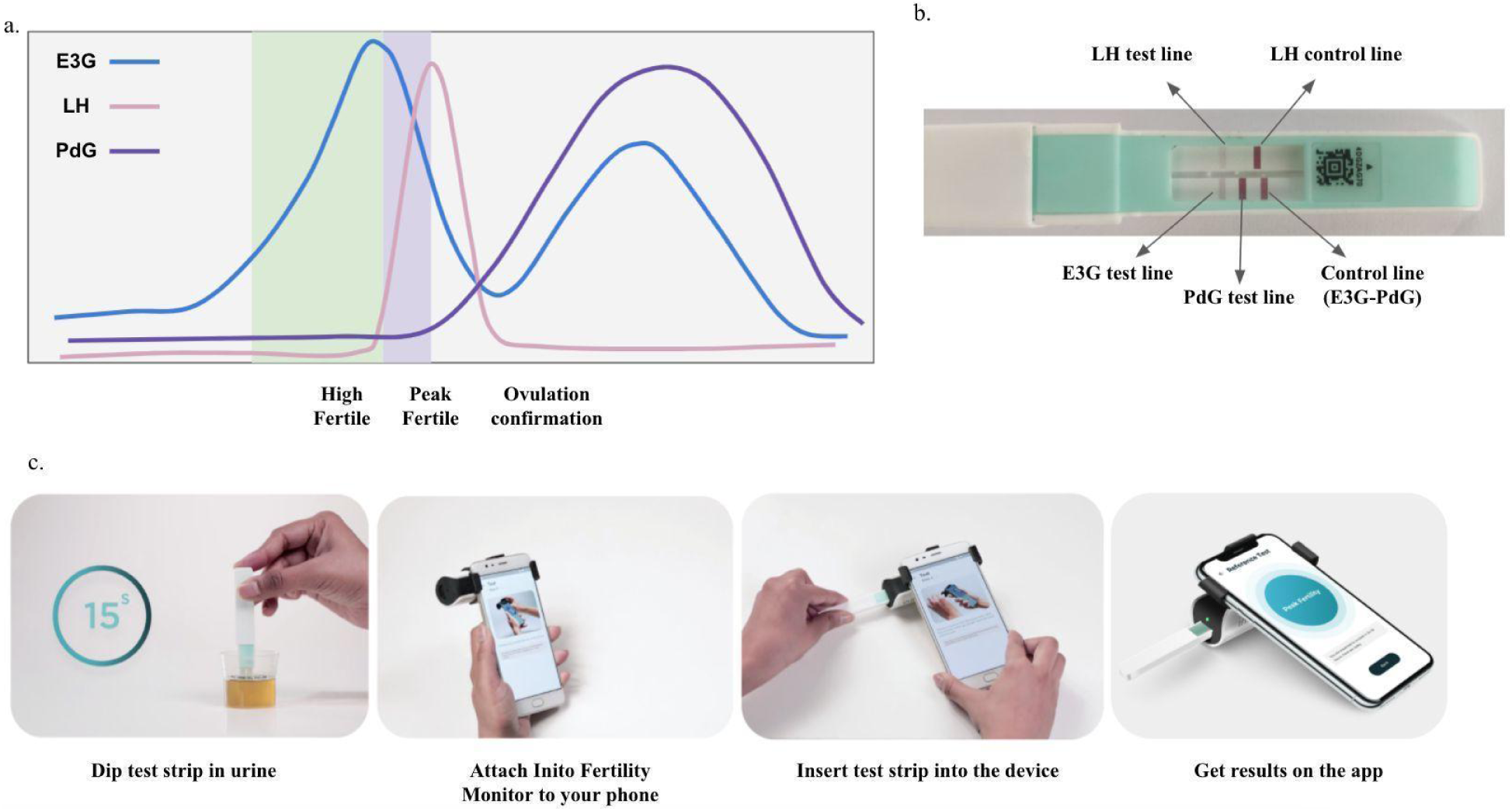
Behaviour of urinary hormones E3G, PdG and LH in a normal menstrual cycle and the prediction of high fertility, peak fertility and ovulation confirmation based on hormone levels (a), location of testing regions in the Inito fertility test strips (b) and schematic of usage of the Inito Fertility Monitor at home (c).

Women use many home-use tests to predict fertile days in the menstrual cycle. Canonical over-the-counter ovulation test kits only assess urinary LH and thus skip a series of fertile days. Other home-use devices available on the market forecast fertile days by tracking LH and E3G. Obtaining a full fertile window can increase the chances of conception by approximately 89%^3^. These home-use techniques, however, have low sensitivity and a narrow detection range. Most of these tests are either visual or provide a binary digital result (positive or negative)^26,27^. However, the hormone patterns are more complex and need quantification. Furthermore, physicians and users are keen on visualizing their hormone trends. Therefore, there is a need for a more sensitive, quantitative home-based test to account for person-to-person and cycle-to-cycle variability while predicting fertile days in women.

We introduce the Inito Fertility Monitor (IFM) (Fig. 1b and c), a mobile mounted, app-connected home-based device that predicts fertile days and confirms ovulation by measuring E3G, LH and PdG simultaneously in urine. The test strip for IFM contains two lateral flow assays: one assay is multiplexed to measure E3G and PdG in a competitive ELISA format, and the other assay measures LH in a sandwich ELISA format. IFM captures the image of the test strip using the Inito mobile application and processes the image to yield the optical density (OD), which corresponds to the concentration of the metabolite. IFM uses a multiscale algorithm to detect the device and eliminate the variations in resolution and aspect ratio due to smartphone variability^11^.

Here, we show that IFM is an effective urinary hormone monitor and has a laboratory-based ELISA-equivalent output that makes it ideal for predicting fertile days and confirming ovulation at home. Using IFM, we could reproduce hormone trends that have been previously reported, for example, premature E3G surge and multiple LH surge in menstrual cycles. In addition, by observing trends in the urinary PdG rise after the LH peak, we propose novel criteria to confirm ovulation earlier in women. Additionally, we present a previously unreported incidence of PdG rise before the LH surge. In conclusion, we suggest that IFM be used by women at home to monitor their fertile days and by clinicians to monitor patients’ hormone profiles and to tailor intervention in the future.

## Materials and methods

### Testing with IFM and repredication from the calibration curve

Test strips were analyzed and qualified using the image processing algorithms and AI algorithms used in Inito Fertility Monitor (Supplementary Method 1). A calibration curve for each batch of Inito Fertility test strips was generated using standard solutions prepared in spiked urine. The ODs obtained from standard solutions were plotted against concentration, and this plot was used to predict concentrations in all further experiments. For testing, the test strips were dipped in the urine samples for 15 seconds. The strips were then inserted into the Inito Fertility Monitor attached to mobile, and values of E3G, PdG and LH were obtained along with the fertility ratings for each day. In the Inito Fertility test strip, E3G and PdG are measured in a competitive ELISA assay format, where the intensity of the respective test lines decreases with increasing concentration, and LH is measured in a sandwich ELISA format, where the intensity of the test line increases with increasing concentration.

### Sample preparation for characterization of Inito Fertility Monitor

Samples used for precision studies, linearity of the reprediction of concentration and cross-reactivity studies were prepared by spiking male urine samples with target concentrations of the metabolites. The male urine samples were tested with ELISA to confirm negligible concentrations of the respective metabolites. Purified metabolites for all studies were obtained from Sigma-Aldrich chemicals as described in Supplementary Table 1.

### Testing with ELISA kits

The same user samples were also used in an ELISA to measure the exact concentration. E3G and PdG were measured using the Arbor Estrone-3-Glucuronide EIA kit (K036-H5) and the Arbor Pregnanediol-3-Glucuronide EIA kit (K037-H5) ELISA kits. Urinary LH was measured using the DRG LH (urine) ELISA kit (EIA-1290). For all runs, solutions of fixed concentration (provided along with the respective kits) were used to generate the standard curve, and the concentration of metabolites in urine samples was calculated from the standard curve generated. All samples were measured in triplicate, and the average value was considered for comparison.

### Study participants

The study design was approved by the Institutional Review Board (IRB) of Sparsh Hospital (EC approval number: CLIN/INI/001). Two groups of women were recruited for the study. The first group of women was shortlisted from a list of students/nurses working in different hospitals near the research facility who volunteered to participate in this trial. The shortlisted participants were further screened and selected based on the exclusion criteria. The second group consisted of women who were looking to try out novel methods for monitoring fertility at home and registered to try Inito at home on the Inito website. Women in the second group were then provided the IFM and the Inito Fertility Test strips to be tested at home. The study protocol was approved by the appropriate institutional review boards (ISRCTN15534557). Informed consent forms were obtained from all volunteers. Women aged between 21-45 years of age were recruited for the study. Women were included in the study if all of the following criteria were met:

1. The age of the participant must be between 21-45 years at the time of registration for the trial.
2. The cycle length of the participants must be between 21-42 days.
3. The cycle length of the participants must not vary by more than +/- 3 days from one menstrual cycle to the other.

Women were excluded if:

a. They have been diagnosed with any infertility conditions and are on infertility medications or hormone replacement therapy containing hCG or LH
b. They were using hormonal contraceptives, including oral, emergency oral, implants, patches, transdermal injections, vaginal ring and progesterone intrauterine systems (IUS)
c. Were consuming clomiphene citrate or other ovulation induction drugs
d. Have recently been pregnant, miscarried or breastfeeding

Reasons for data exclusion were irregularities in testing leading to insufficient data points for any conclusion and failure to meet the selection criteria. The characteristics of those who were considered for the study are summarized in Table 1.

**Table 1.** Characteristics of eligible participants for the study

|  | Group I (samples collected directly)<br>(n=100) | Group II (Using IFM at home)<br>(n=52) |
| --- | --- | --- |
| Age (y), mean (SD) | 27.3 (4.6) | 27.6 (4.1) |
| Cycle length (d), mean (SD) | 30.4 (6.3) | 27.6 (5.4) |
| BMI |  |  |
| <18.5, n(%) | 21 (21) | 0 (0) |
| 18.5-24.9, n(%) | 66 (66) | 41 (78.9) |
| 25-29.9, n(%) | 10 (10) | 5 (9.6) |
| >30, n(%) | 3 (3) | 6 (11.5) |
| Trying to conceive, n(%) | 31 (31) | 52 (100) |
| Time trying to conceive (cycles), n(%) | <2, 22 (70.9%)<br>2-4, 9 (29.1%) | <2, 10 (19.2%)<br>2-4, 12 (23.1%)<br>4-6, 30 (57.7%) |

### Research Ethics

The study protocol was approved by the Institutional Review Board at Sparsh Hospital, Bangalore. The approval number assigned was CLIN/INI/001. Informed consent forms were obtained from all participants before being enrolled in the study, and all studies were carried out in accordance with the Declaration of Helsinki.

### Study design

Urine samples collected from women in the first group were frozen, transported to the testing site and tested with the IFM on the same day. Freeze-thaw cycles have been previously shown to have no effect on the concentration of urinary hormones^12,13^. For the second group, IFM and Inito Fertility test strips were sent to participants to perform the test at home. Standard instructions for use were provided with the IFM box and the test strips. All participants had the option to contact the manufacturer’s helpline number for any additional information concerning the IFM. Women began using the IFM at the start of the menstrual cycle after admission into the study. Participants were instructed to take tests on specific cycle days when instructed by the application. Hormone data from all participants were collected via the Inito application. Although women were recruited for one cycle each, participants were excluded if they did not take tests on the required days. The study design is summarized in Figure 2.

**Figure 2:**
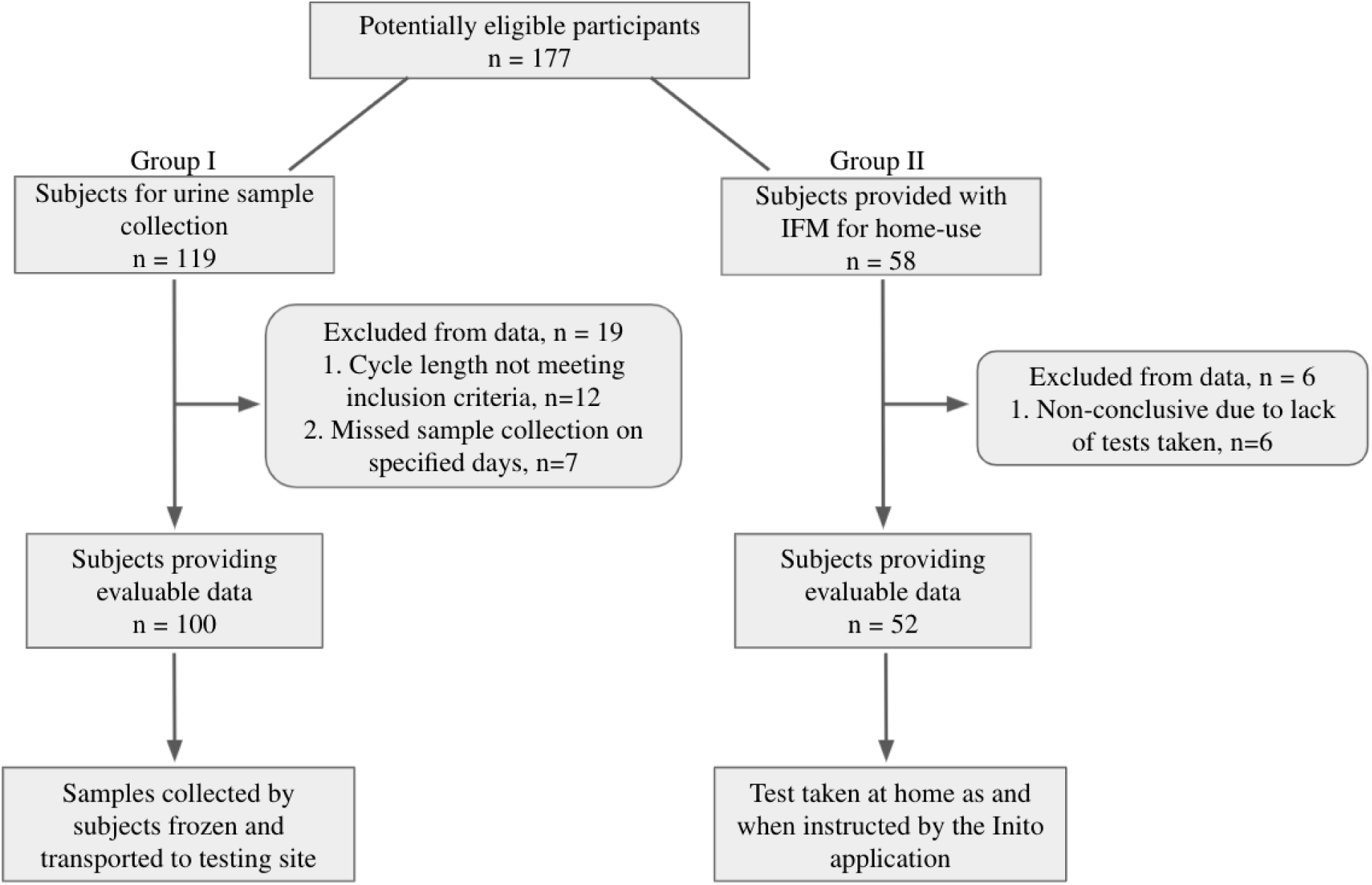
Flow of participants through the study

### Definitions

#### Cycle day 1 is defined as the day of bleeding

The day of ovulation is the day of the peak LH value. An LH surge event is defined as any day where the LH value exceeds 5 mIU/ml from the baseline value in urine. A cycle with multiple LH surges is any cycle that has smaller LH surges before the actual LH peak.

A normal E3G surge is the rise in E3G expected to be seen at least 5-6 days before the LH peak value, and any surge before that is considered to be premature.

To confirm ovulation, the canonical method implements measurement of urinary PdG at 7 days after the LH peak. If the urinary PdG value obtained was greater than 5 µg/ml, the cycle was classified as ovulatory^16,17^.

## Results

### Inito fertility monitor accurately predicts the concentration of urine metabolites and is specific to the metabolites of interest

The accuracy of the Inito Fertility test strips was assessed by reprediction of the concentrations of E3G, PdG and LH in standard solutions prepared in male urine not containing any of the metabolites. Six spiked solutions containing all three metabolites were prepared from the respective stock solutions as per Supplementary Table 2. Each solution was tested with five different Inito fertility test strips, and the recovery percentage was calculated. We found that the average recovery percentage across the five tests was 100.16% for E3G, 104.63% for PdG and 104.74% for LH. The observed concentration obtained from IFM was also highly correlated to the expected concentration in the spiked male urine (Fig. 3a-c), implying a high accuracy in predicting the concentrations.

**Figure 3:**
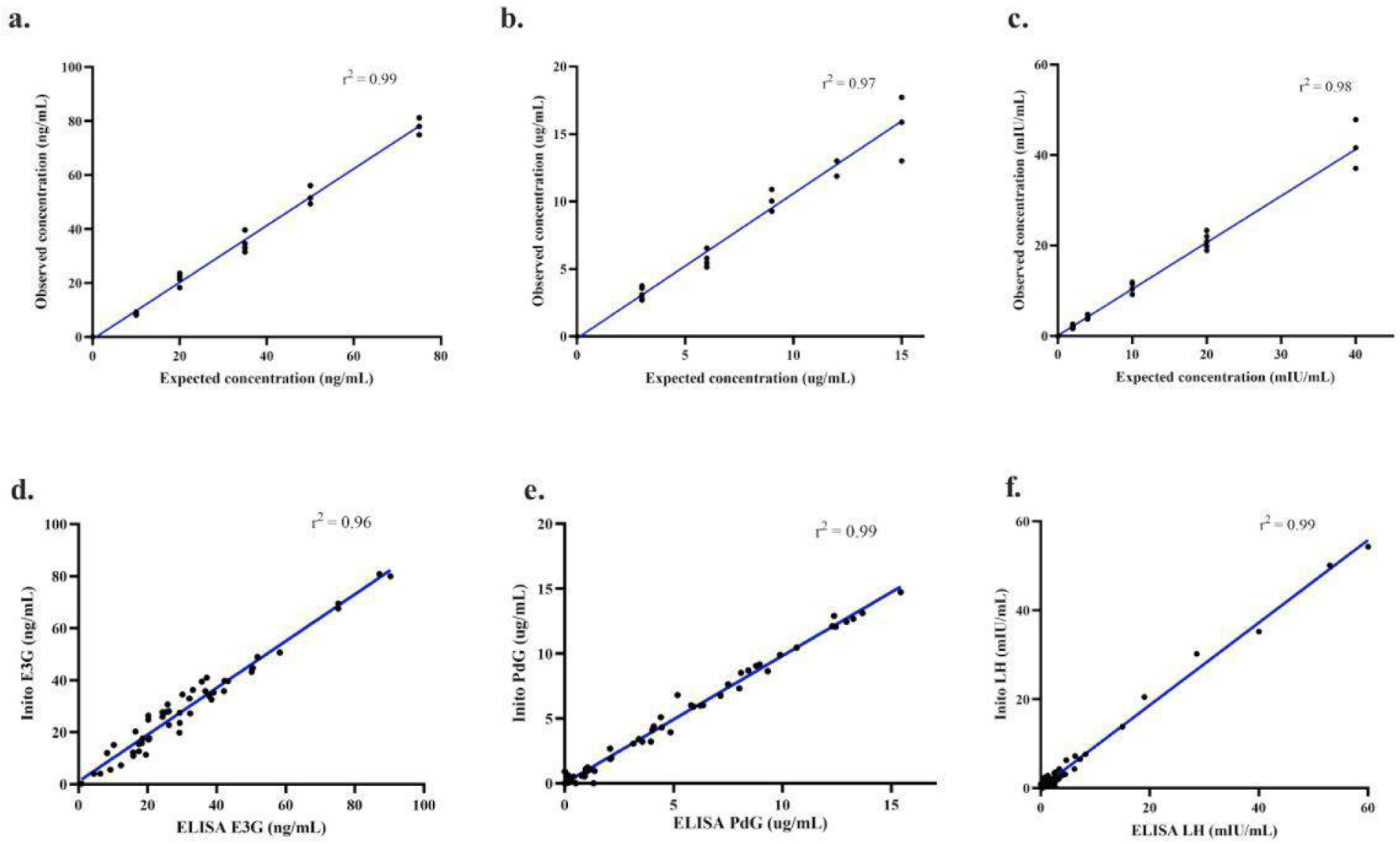
Correlation between observed concentrations obtained from IFM and expected concentrations of E3G (a), PdG (b) and LH (c). Linear regression (blue) for the comparison between re-predicted hormone values from urine samples obtained using IFM and ELISA: E3G (d), PdG (e) and LH (f).

Since the Inito Fertility test strips are manufactured in a batchwise fashion, we wanted to ensure that the accuracy is maintained across lots. Therefore, ten strips from different lots were tested with four different concentrations of E3G, PdG and LH, and the coefficient of variation (CV) was calculated. All three assays were found to have a CV of less than 10% across different lots (Supplementary Table 3), indicating that the recovery percentage does not vary significantly from test strip to test strip. Therefore, IFM is an accurate device to measure urinary hormones and is consistent from test to test.

Next, we wanted to analyse the specificity of Inito fertility test strips towards the metabolites of interest. We identified the components of human urine that could potentially interfere with the test and spiked negative male urine with these compounds. The concentration of each metabolite was selected to encompass normal and pathological conditions. We tested four strips from different batches with the spiked male urine and found that the test strips did not cross-react with any urine metabolite at even the highest possible physiological condition (Supplementary Table 4). This suggests that the Inito fertility test strips are specific to the metabolites of interest only.

### Urinary hormone concentrations estimated by IFM and ELISA are strongly correlated

Enzyme-linked immunosorbent assay (ELISA) is a standard laboratory procedure for measuring concentrations of chemical compounds accurately. Therefore, we wanted to compare the efficacy of estimation by Inito with respect to ELISA. For this, we chose all the urine samples collected by 3 random women (out of 100) over one complete cycle. The cycle lengths of these three women were 26 days, 32 days and 22 days, accounting for a total of 80 samples that were tested with both Inito and ELISA, and the correlation was calculated. We found that the results obtained from ELISA and Inito are highly similar (r^2^=0.96 for E3G, r^2^=0.99 for PdG, and r^2^=0.99 for LH) and are linearly correlated (Fig. 3d-f), implying that measurement by Inito is almost as accurate as measurement by a standard laboratory technique.

### The incidence of premature E3G surge and multiphasic LH surge may be higher in women than expected

Our first idea was to observe how well the 6-day high fertile window predicted by the IFM correlates with the occurrence of ovulatory cycles. We observed that on average, a 6-day fertile window correlates the best with the occurrence of an ovulatory cycle compared to lower fertile days or higher (Supplementary Figure 2).

Our study design allowed us to monitor urinary E3G, LH and PdG on all days of the follicular phase and the luteal phase, including the day of ovulation. Therefore, we decided to observe the occurrence of aberrant E3G and LH patterns in the volunteer menstrual cycles. Specifically, we investigated the prevalence of premature E3G surge and multiphasic LH surge (multiple LH peaks). We observed that among the cycles that displayed LH rise and ovulation (confirmed using urinary PdG), 39 cycles recorded multiple surges of LH before the actual peak that caused ovulation, and 16 cycles recorded a single dominant LH surge. Thirty-nine cycles were anovulatory and had no LH surge. However, 6 cycles were anovulatory despite a single surge of LH, a phenomenon previously described as luteinized anovulation. While the occurrence of multiphasic LH is coincident with previous studies^14,15^, the percentage of cycles with a single LH surge leading to ovulation may actually be lower than what had been previously assumed. With respect to E3G, we observed that 20 cycles showed a significantly high E3G in the early part of the cycle, typically more than 6 days before the LH peak and before the expected E3G surge (right before LH surge), the occurrence of which is coincident, albeit slightly higher than previously reported studies^15^.

### Monitoring the continuous rise of urinary PdG after LH surge provides early confirmation of ovulation

The serum progesterone (P4) concentration is widely considered a confirmatory marker for ovulation. Typically, a mid-luteal phase value of >3 ng/mL was used to confirm ovulation. Recently, it has been shown that mid-luteal phase measurement of urinary PdG (a threshold value of >5 µg/mL) has a good correlation with serum P4 behaviour and can also be used to confirm ovulation^16,17^. However, both methods require the users/patients to wait for almost 7 days after the LH peak value.

For our analysis, to classify a menstrual cycle as ovulatory or anovulatory, we selected the same urinary PdG threshold values as previously discussed, and this was regarded as the actual positive or negative value for further evaluation of the novel criteria. The baseline urinary PdG was determined before the LH surge as the average of the PdG levels and compared with the PdG levels 2-3 days after the LH surge. We found that the distribution of fold change in ovulatory cycles was substantially different from the fold change observed in the anovulatory cycles during the first three days after the LH peak (Fig. 4a and b). From this observation, we inquired if measuring a particular fold rise within 3 days after the LH peak could be used as a criterion to predict ovulation earlier. We decided to judge these criteria by performing an ROC analysis. We found that a fold change of 2.75x within 3 days could accurately distinguish ovulatory from anovulatory cycles. The criteria had a specificity and sensitivity of 100% in detecting ovulation and were significant in differentiating ovulatory from anovulatory cycles (p<0.0001, AUC: 0.981, Fig. 4c and Table 2a). In addition, we applied this criterion to the set of users who used the IFM at home to predict ovulation. We found that the new criteria could differentiate the 18 ovulatory cycles from the 4 anovulatory cycles with 100% precision (Table 2b). Our findings show that constant urinary PdG monitoring can provide earlier ovulation confirmation than a single PdG/P4 measurement in the mid-luteal phase.

**Figure 4:**
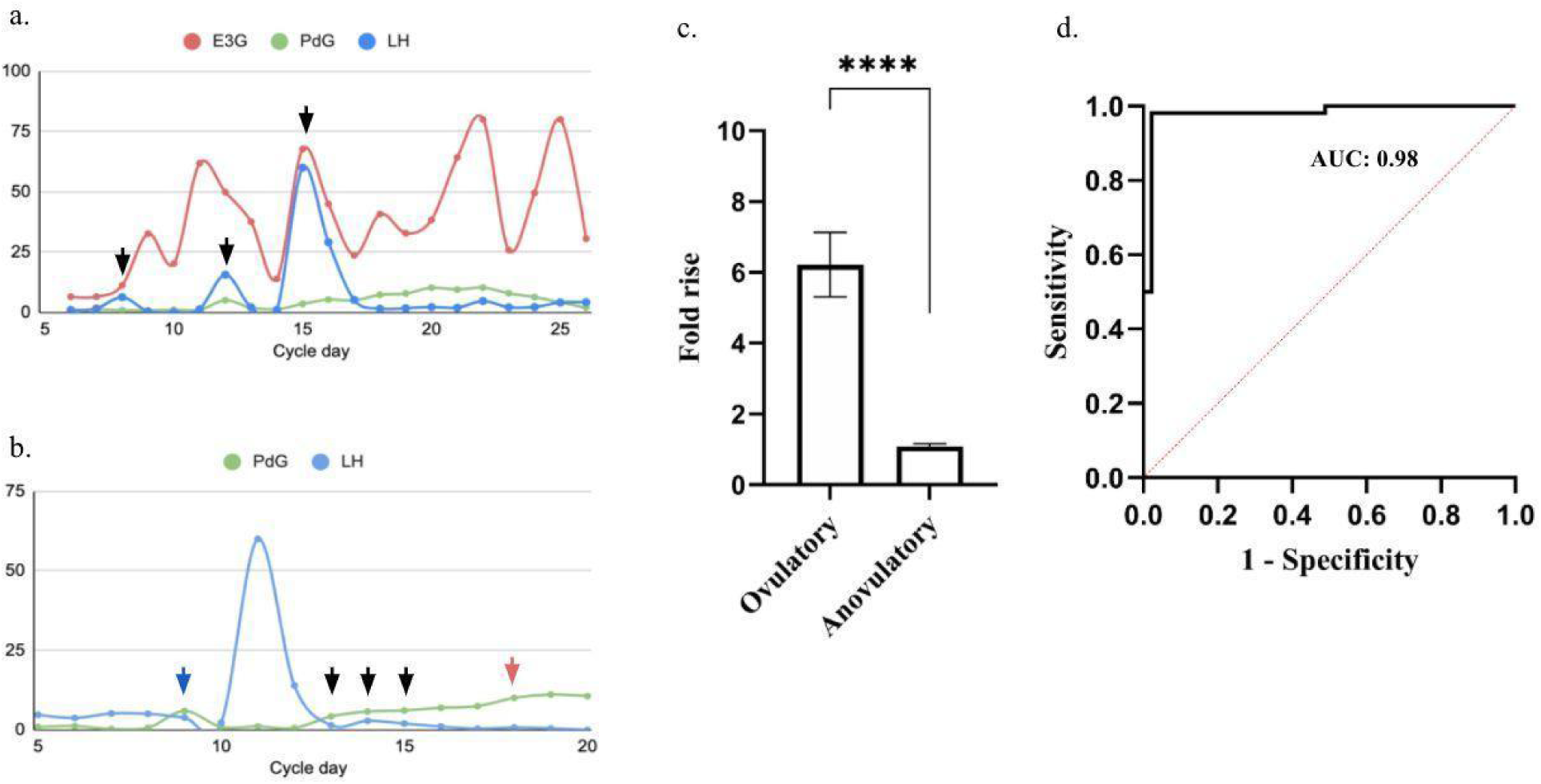
a. Representative hormone chart for multiphase LH surges with each event of LH surge marked (black arrows), b. Representative graph showing PdG rise before the LH surge (blue arrow) and the gradual rise in PdG after LH peak (black arrow) for confirming ovulation as opposed to a single test mid-luteal phase (red arrow), c. Comparison of fold rise in PdG levels within 3 days after LH surge in ovulatory versus anovulatory cycles (****p<0.0001 and d. ROC analysis to evaluate the accuracy of the proposed criteria to predict ovulatory and anovulatory cycles.

**Table 2.** Decision matrices comparing the outcomes of prediction of ovulation between the existing criteria (mid-luteal phase measurement) and proposed criteria (criteria proposed using IFM) applied to Group I (a) and Group II (b). Positive and negative indicate ovulatory and anovulatory, respectively.

|  | **Positive**<br>**(existing**<br>**criteria)** | **Negative**<br>**(existing**<br>**criteria)** |
| --- | --- | --- |
| **Positive**<br>**(proposed**<br>**criteria)** | 55 | 0 |
| **Negative**<br>**(proposed**<br>**criteria)** | 0 | 45 |

Table 2. Decision matrices comparing the outcomes of prediction of ovulation between the existing criteria (mid-luteal phase measurement) and proposed criteria (criteria proposed using IFM) applied to Group I (a) and Group II (b).
|  | **Positive**<br>**(existing**<br>**criteria)** | **Negative**<br>**(existing**<br>**criteria)** |
| --- | --- | --- |
| **Positive**<br>**(proposed**<br>**criteria)** | 18 | 0 |
| **Negative**<br>**(proposed**<br>**criteria)** | 0 | 4 |

### Luteinizing hormone peak is preceded by a distinct rise in urinary PdG

A rise in estradiol levels prior to the LH surge is observed in canonical menstrual cycles. Progesterone, however, is thought to be the baseline during the follicular phase (Fig. 1a). Our study design allowed us to monitor urinary PdG prior to LH surge as well. Furthermore, the sensitivity of the Inito Fertility test strips allowed the smallest changes in the urinary PdG to be visualized. We observed that urinary PdG increased in Group I prior to the LH surge by an average of 3.16+/-0.7-fold in 52/55 cycles from the baseline stage (in cycles where there was a prominent LH surge). This surge was found to occur on average 2.87+/-0.9 days before the day on which LH peaked. Only 2 cycles illustrated this phenomenon in the 6 anovulatory cycles of Group I, where there was an LH surge. In Group II, we found the frequency of this phenomenon in 43/52 cycles by re-evaluating this observation. Although we remain unsure of the source of this urinary PdG, the phenomenon appears to be highly conserved during menstrual cycles.

## Discussion

Optimal intercourse timing has been established as a key parameter influencing the frequency of pregnancy in women^18^. Previous studies have addressed the existence of a 6-day fertile window as the optimum fertile window correlated with an increased probability of pregnancy^19,20^. In addition, the fertile window prediction is well correlated with the concentrations of urinary hormones, specifically the growth of E3G and LH. Nonetheless, approximately 26-37 percent of natural cycles are anovulatory^8^ and may occur despite normal hormonal conduct, for instance, in the case of luteinized unruptured follicles ^21,22^. Thus, to ensure ovulation, the estimation of the fertile window is not adequate and therefore may not be well associated with pregnancy results.

The Inito Fertility Monitor (IFM) is a home-based diagnostic device that measures urinary E3G, PdG and LH concentrations. Measurement of these three hormones enables confirmation of ovulation after prediction of a potential fertile window for women. The performance analysis of IFM proves that it is a very sensitive, accurate and laboratory equivalent device for monitoring the urinary hormone profiles of women.

From a population analysis of women who volunteered to collect urine over one menstrual cycle and women who used IFM at their homes, our system classified 40.2% of the menstrual cycles as anovulatory in the random trial we conducted, which is similar albeit slightly higher than what has been observed in previous studies^8^. Since this study was performed in a different group of women compared to the previous study (different populations), it can be inferred that the severity of this subfertility class may be similar across different populations, and hence, a generic regime can be implemented for different women assuming that the extent of this problem is the same.

The occasional cases of luteinized anovulation could also be captured, the frequency of which often coincides with what was previously recorded^22^. From the hormone data obtained, we propose a novel paradigm for faster confirmation of ovulation by virtue of continuous monitoring of urinary PdG. This paradigm could encompass all cycles that were classified as ovulatory according to the existing thresholds and methods of confirmation proving that the measurement of mid-luteal phase progesterone may be redundant and that the rise in progesterone could be captured much earlier to confirm ovulation. Since pre-implantation intercourse diminishes fecundability by almost 26%^29^, application of this method to confirm ovulation may help in increasing the fecundability since women can decide not to have intercourse once they receive a confirmation of ovulation.

From the hormone trends, we also show that urinary PdG rises in a number of cycles before the LH surge. Previously, the occurrence of a preovulatory progesterone rise was reported by Hoff et al.^23^ The role of this preovulatory progesterone has been proposed to be triggering the LH surge, and a premature rise in this has been indicated in the pathology of polycystic ovarian syndrome (PCOS)^24,25^. The source of this progesterone has been hypothesized to be follicular, and the surge is supposed to occur 12-24 hours before the expected LH surge. The progesterone surge observed by us occurs much prior to this period and hence may be a separate phenomenon possibly correlated to a particular developmental phase of the follicle. However, since the occurrence is highly conserved, it is likely that this feature may be an indicator of a common feature associated with normal natural cycles and could help in differentiating between other infertility-related conditions.

## Conclusion

The results from this study indicate that IFM is an accurate home-based fertility monitor to help women determine their fertile window as well as to confirm ovulation. Since the Inito Fertility Monitor provides the hormone charts to the users, they can monitor the effects of changes in lifestyle on their hormones and select regimes that contribute to the best hormone behavior.

While IFM is currently designed for use at home, the sensitivity and repeatability of results allow it to be used as a device for basic understanding of hormone trends in women as well^28^. This suggests that in the future, this platform could be used by physicians as well for monitoring the hormone profiles of their patients. A remote monitoring platform such as this would make the path to diagnosis and treatment of subfertility conditions faster since women can use it at home, obtain hormone charts and share them with their doctors. In addition, using IFM, doctors could also visualize the effects of treatments on their patients and modify them to obtain the best possible output in terms of fecundity.

Additionally, more clinical trials involving women using IFM at home can open avenues of the device’s use in handling cycle-related factors such as PMS and monitoring pregnancy health. Furthermore, monitoring hormone patterns of specific groups of infertile women can aid in formulating algorithms that can help in a basic prognosis of these infertility conditions at home.

## Data Availability

All the data is available in the manuscript. Any additional data will be made available upon inquiry.

## Author contributions

S.P. and V.A. designed the study. S.P. planned the study procedure and conducted the clinical trials. S.P. and D.D. performed the laboratory analysis and established equivalence with ELISA. S.P. analyzed the data. S.P. and V.A. analyzed user hormone trends. S.P. wrote the manuscript. S.P. and V.A. V were involved in editing the manuscript.

